# Acceptance of and preference for COVID-19 vaccination in healthcare workers: a comparative analysis and discrete choice experiment

**DOI:** 10.1101/2020.04.09.20060103

**Authors:** Chuanxi Fu, Zheng wei, Fengchang Zhu, Sen Pei, Shunping Li, Liuren Zhang, Xiaohui Sun, Yue Wu, Ping Liu, Mark Jit

## Abstract

**Background:** A major obstacle to successful coronavirus disease (COVID-19) vaccine rollout is vaccine hesitancy. Acceptance of and preferences for COVID-19 vaccination among healthcare workers (HCWs) is critical, because they are a key target group for vaccination programs, and they are also highly influential to vaccine uptake in the wider population. This study sought to comparatively determine the acceptance of and preference for COVID-19 vaccination among HCWs and the general population.

**Methods:** An Internet-based, region-stratified discrete-choice experiment was conducted among 352 HCWs and 189 general population respondents recruited in March 2020 from 26 Chinese provinces. We accessed knowledge of disease, attitude towards and acceptance of COVID-19 vaccination. Several attributes (related to COVID-19 disease, COVID-19 vaccination and one social acceptance) were identified as key determinants of vaccine acceptance.

**Results:** HCWs had a more in-depth understanding of COVID-19 and showed a higher willingness to accept COVID-19 vaccines with lower effectiveness and/or more severe adverse effects compared to the general population. 76.4% of HCWs (vs 72.5% of the general population) expressed willingness to receive vaccination (χ^2^=2.904, *p*=0.234). High levels of willingness to accept influenza (65.3%) and pneumococcal (55.7%) vaccination were also seen in HCWs. Future COVID-19 disease incidence (OR: 4.367, 95% CI 3.721–5.126), decisions about vaccination among social contacts of respondents (OR 0.398, 95% CI 0.339–0.467), and infection risk >30% (OR 2.706, 95% CI 1.776–2.425) significantly increased the probability of vaccination acceptance in HCWs.

**Conclusion:** Multi-component interventions to address the key determinants of hesitancy in both HCWs and in the general population should be considered for COVID-19 vaccination programs.

## 1. Introduction

The coronavirus disease (COVID-19) caused by the severe acute respiratory syndrome coronavirus 2 (SARS-CoV-2) was initially identified in December 2019 in Wuhan, a city in Central China, and soon evolved into a global pandemic, with 173 million cases and 3.74 million deaths reported as of June 9, 2021, as well as substantial economic losses^1-3^. Interventions such as physical distancing and quarantine have slowed viral spread and contributed to flattening of the epidemic curve. However, the COVID-19 epidemic is unlikely to subside in the absence of continued physical distancing measures until large proportions of populations have been immunized either through recovery from infection or following vaccination. At least 287 research programs to develop vaccine candidates against SARS-CoV-2 have been launched since 2020. Many of these candidates have been successful in clinical trials, and are being used in populations around the world^4^.

However, a major obstacle to successful COVID-19 vaccine rollout is vaccine hesitancy. COVID-19 vaccine hesitancy can be viewed from the lens of the Increasing Vaccination Model, a behavioral science framework to understand people’s decision to get vaccinated ^5^. Practical factors such as vaccine supply can also affect their ability to act on the motivation to get vaccinated^6, 7^.

A key target group for COVID-19 vaccination is healthcare workers (HCWs). The World Health Organization recommends HCWs, especially those at high risk of acquiring and transmitting SARS-CoV-2, as a key priority group for vaccination^8^. HCWs are generally at higher risk of getting infected by various pathogens such as influenza viruses and SARS-CoV-2 than the general population ^9, 10^. They are also vital to communicating information about vaccination to potential vaccinees and their caregivers. The importance of vaccine recommendations made by HCWs to potential vaccinees in the decision-making process has been well documented: HCWs have been found to be one of the strongest influencers in vaccination decisions ^11-13^.

In this study, we investigated the knowledge of, attitudes toward, and preferences for future COVID-19 vaccination among HCWs in comparison with the general public, in order to inform measures to improve vaccine acceptance and coverage in general populations. We used discrete choice experiments (DCEs) among both HCWs and the general population. DCEs, initially used in marketing research, have been widely used to assess preferences for medical intervention such as vaccination programmes. In a DCE, the intervention is described using different attributes, such as vaccine efficacy and cost, with each attribute measured using a number of discrete attribute levels (e.g. vaccination cost of $10, $20, or $30). The DCE presents respondents with a series of choice sets containing collections of attribute levels, and the preferences that respondents have for one choice set over another indicates the relative importance of the attributes and attribute levels on their decision preferences ^14^. DCEs have been used to explore vaccination preferences in different population groups, such as girls and older adults ^15-17^.

## 2. Materials and methods

### 2.1 Participants and design

The study enrolled HCWs aged 20–59 years from hospitals, local Centers for Disease Control and Prevention (CDC), and health community centers. Based on the total number of confirmed COVID-19 cases reported until March 15, 2020 ^18^, we divided all provinces in mainland China into two categories: (i) provinces with high cumulative COVID-19 incidence (exceeding 8 cases per 100,000), including Hubei, Guangdong, Zhejiang, and Henan, and (ii) provinces with low cumulative incidence (at or below 8 cases per 100,000).

On March 17 and 18, 2020, following the cessation of large epidemics in Hubei province and other parts of China, we initiated an online survey via WeChat, China’s most popular messaging app that has a monthly user base of more than 1 billion people. Two rounds of Delphi and preliminary experiment were conducted before formal survey. We first enrolled 20 HCWs, who then invited their WeChat colleagues or contacts (30 to 40 subjects) to the survey from both high and low cumulative incidence provinces to participate in the survey by sending them a WeChat link. To compare how COVID-19 vaccination acceptance differed among HCWs compared to the general population, we also invited adult participants from non-health care backgrounds, recruiting a sample that was approximately half the size of the HCW sample. We powered the study to estimate a COVID-19 vaccination acceptance rate of 65% in both groups, with a standard error of 0.05 in the HCW group and 0.08 in the general population group, and with 5% significance. This is based on the acceptance level of H1N1 pandemic vaccine (64%) or seasonal influenza vaccine (64%) measured in August 2009^19^. Using this, we estimated that 348 HCW participants (σ=0.05, α=0.05) and 136 general population participants (σ=0.08, α=0.05) were required. The study approval was obtained from the Institutional Review Board of Zhejiang Chinese Medical University (ZCMU) and anonymity was guaranteed to participants.

### 2.2 Data collection

The survey was conducted through a self-administered, anonymous questionnaire, which comprised of five sections: (i) demography; (ii) knowledge of and attitudes about SARS-CoV-2 infection, including susceptibility, health outcomes, subgroups at high risk of mortality, effective treatment, virus mutation, future epidemic trends and infection risk; (iii) acceptability of COVID-19 vaccination, including their opinion about the necessity of getting vaccinated, time before the vaccine is likely to become available, population groups most in need of vaccination, desire to receive vaccination, minimum level of vaccine effectiveness, most serious adverse effects following vaccination, highest number of doses needed, highest acceptable cost, and confidence in domestically manufactured vaccines; (iv) post-epidemic behavior, including intention to receive seasonal influenza vaccination or pneumococcal vaccination, and maintenance of other (unspecified) protective measures; and (v) preference about receiving COVID-19 vaccination (eSupplement1). All questions in the survey were designed based on evidence in the literature about the determinants of people’s decisions to get vaccinated^2, 3, 7, 10^. The SO JUMP software (Ranxing Info Technology Co., Ltd), an online survey platform, was used for the online survey.

### 2.3 Discrete choice experiment

#### 2.3.1 Attributes and hypothetical profiles

To access the preferences for COVID-19 vaccination among HCWs and the general population (i.e. section (v) of the survey described in Section 2.2), we used a discrete choice experiment (DCE) approach. We selected the most relevant COVID-19 vaccination attributes and their levels based on a literature review, group discussions with five HCWs and five individuals from the general population, and expert interviews of those in the field of vaccines and epidemiology. The levels of each attribute were specified to ensure that the thresholds were adequate and feasible to capture the interviewees’ attention. Seven attributes were finally identified as the key determinants of vaccination decisions, including: three disease-relevant attributes (probability of infection, severity and probability of death once infected, and future epidemic trends), three vaccine-relevant attributes (vaccine efficacy, vaccine safety, and out-of-pocket vaccination cost), and one attribute of social acceptance (Table 1)(eSupplement2). Each respondent was assigned a survey consisting of 16 hypothetical profiles, each containing a specific level for each attribute. The 16 profiles were derived from 648 (3×2×3×2×2×3×3) candidate attribute profiles, and determined using a fractional factorial design based on orthogonal arrays (ORTHOPLAN procedure, IBM SPSS Statistics). Then, the selected profiles were randomly distributed between eight choice sets, each of which was comprised of two hypothetical profiles (Scenario A and Scenario B). For each choice set, participants were invited to select their preferred COVID-19 vaccination option from either Scenario A or B(eSupplement1).

**Table 1.** Attributes and levels included in the discrete choice experiment

| Attributes | Levels |
| --- | --- |
| Infection probability | 1–14%<br>15–30% ($\beta_1$)<br>>30% ($\beta_2$) |
| Case-fatality ratio | <3%<br>3–15% ($\beta_3$) |
| Possible trends of the epidemic | Disappears in summer, similar to SARS<br>Seasonal epidemic, similar to influenza ($\beta_4$)<br>Nonchronic, similar to tuberculosis ($\beta_5$) |
| Vaccine safety | Safe, with mild side effects ($\beta_6$)<br>Uncertain, it is a new vaccine |
| Vaccine efficacy | 50–80%<br>>80% ($\beta_7$) |
| Out of pocket of the vaccination | Free<br>RMB 100 (US\$14.3) ($\beta_8$)<br>RMB 300 (US\$42.9) ($\beta_9$) |
| Acceptance of social contacts | Neutral/no opinion ($\beta_{10}$)<br>Refusal ($\beta_{11}$)<br>Encouragement |

#### 2.3.2 Sensitivity analysis

An additional choice set was assigned to each participant to identify participants who made inconsistent choices, indicating that they may be unable to understand the choices being presented. In this set, the attributes assigned to Scenario A were all better than those assigned to Scenario B. Participants who “failed” in this test (choosing Scenario B over Scenario A) were excluded from the main analysis although they were still included in a sensitivity analysis.

### 2.4 Data statistics

We did not run through mixed logit model in that most attributes’ levels were qualitative variables. To estimate the influence of each attribute in the DCE on participants’ preferences about vaccination, we used a binary logistic regression model shown in the equation below. This assesses the preference weight (odds ratio [OR]) for each attribute level to the probability of a participant preferring to be vaccinated. Here, β_1–11_ are coefficients corresponding to each of the attributes in the DCE; X_i_ is a vector of alternative levels of the attributes selected by each individual; β_0_ is a constant term, and Error indicates a random term following a type I extreme value distribution (Table 1).

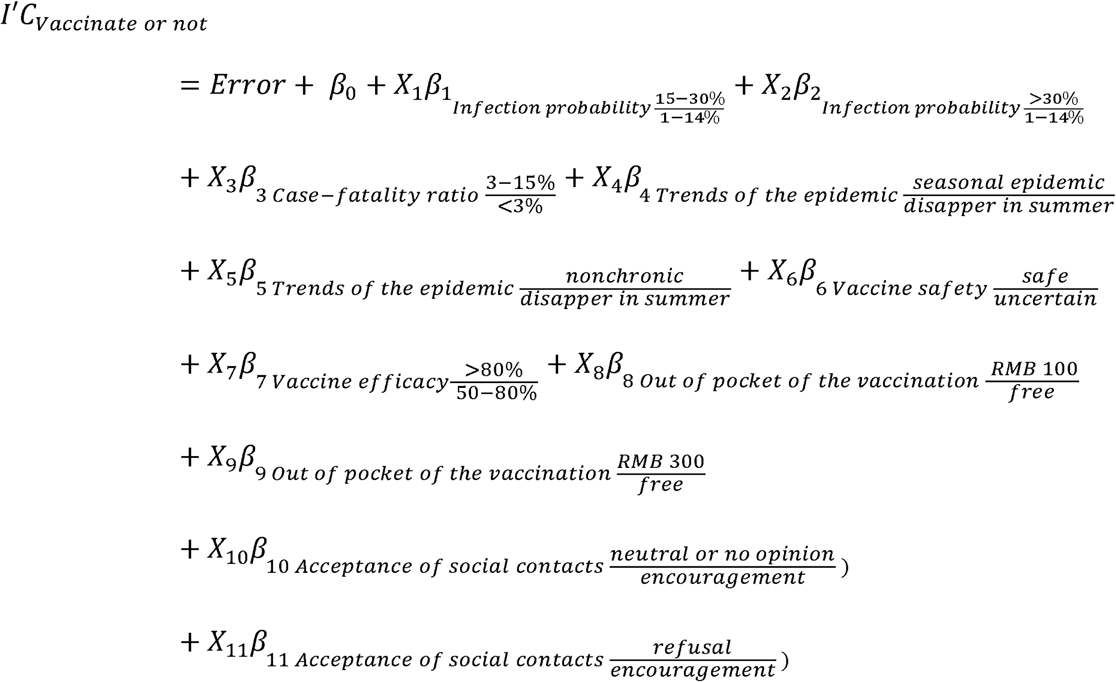

A descriptive analysis of respondents’ knowledge of, attitude about, and acceptance of COVID-19 vaccination was conducted using SPSS Version 25.0 (IBM Corporation, New York, NY, United States). The statistical analysis of the DCE results was carried out in STATA 16 (StataCorp LLC, Texas, United States).

## 3. Results

### 3.1 Demographic characteristics

We approached 583 individuals to complete the survey. Of these, 22 (3.85%) did not complete the survey, and 20 (3.56%) were excluded based on our selection criteria (13 were living outside China, and 7 did not fulfil the age criteria of being 20-59 years old). Of the 541 participants selected for the analysis, 352 (65%) were HCWs and 189 (35%) were general population respondents, from 26 Chinese provinces.

Approximately 303 (56%) of the respondents were from high cumulative incidence provinces, and 70 (23%) were from Wuhan. Nearly 60% (207/352) of the HCWs were female, and 69% (242/352) were younger than 40 years. There were no significant differences between HCW and general population respondents in terms of sex (χ^2^=0.719, *p*=0.396) or age (χ^2^=4.834, *p*=0.184). Approximately 90% (313/352) of HCWs had a bachelor’s degree or higher qualification and were better educated than the general population sample (59.3%, 112/189; χ^2^=65.057, *p*<0.001; Table 2).

**Table 2.**
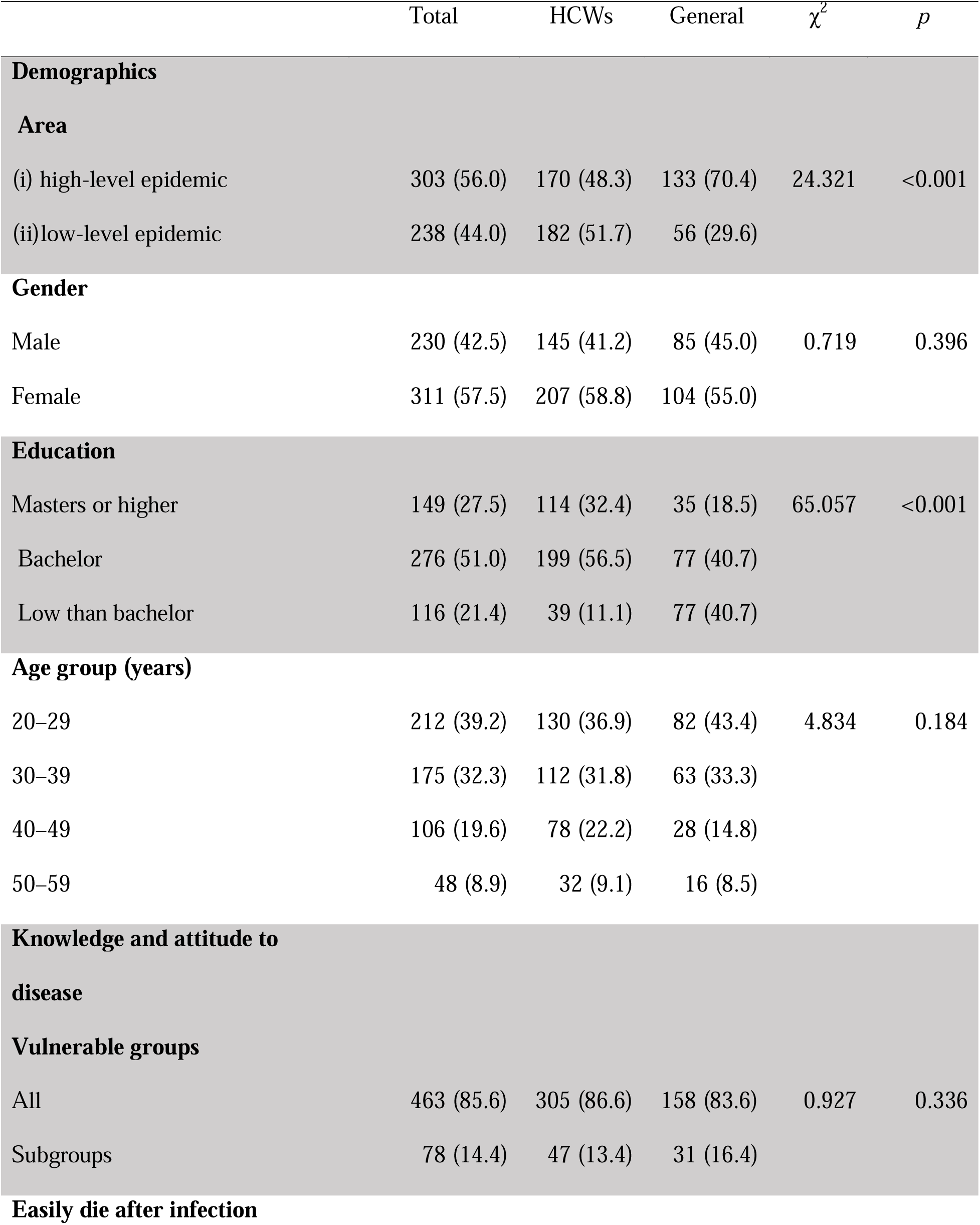
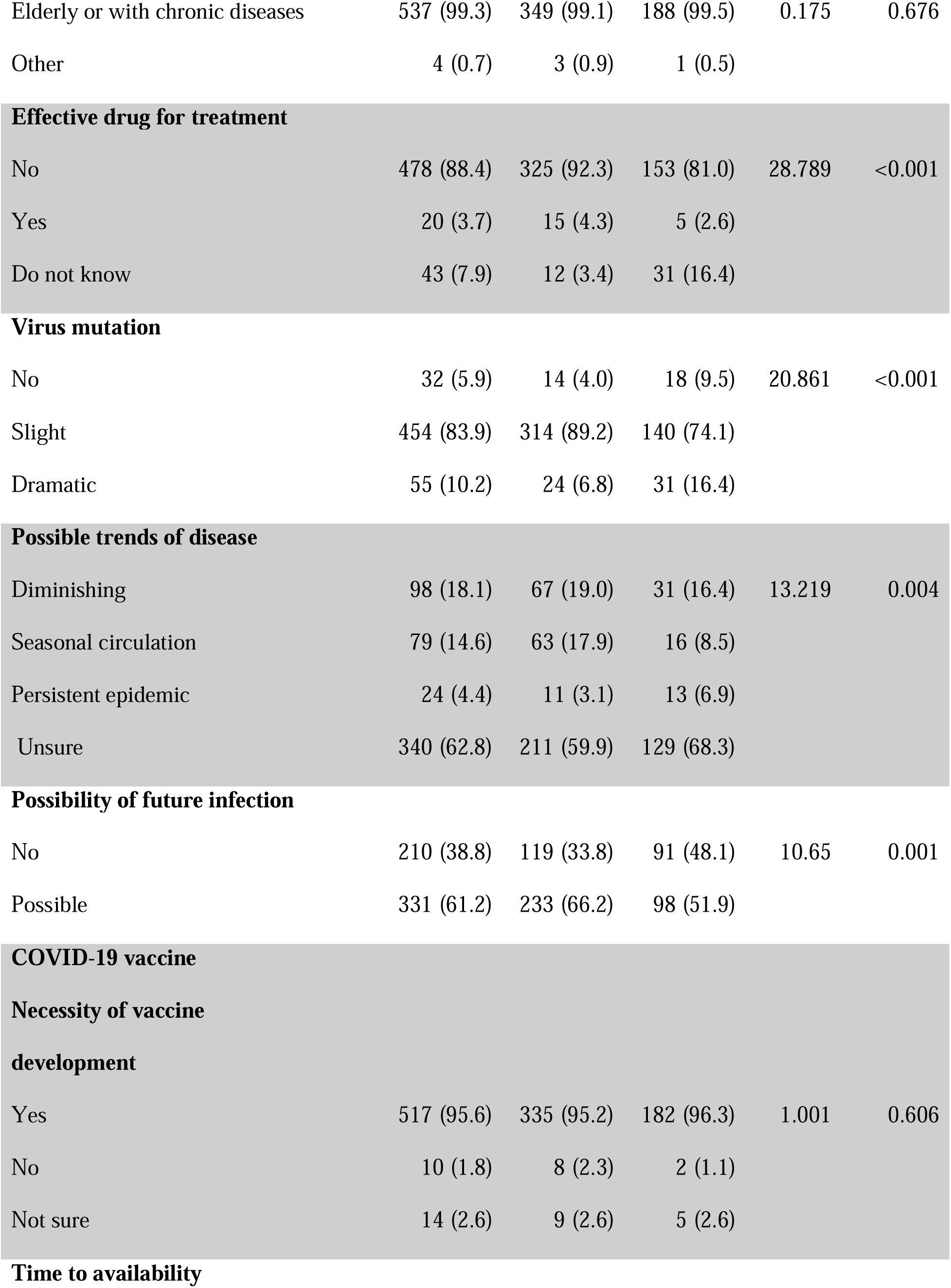
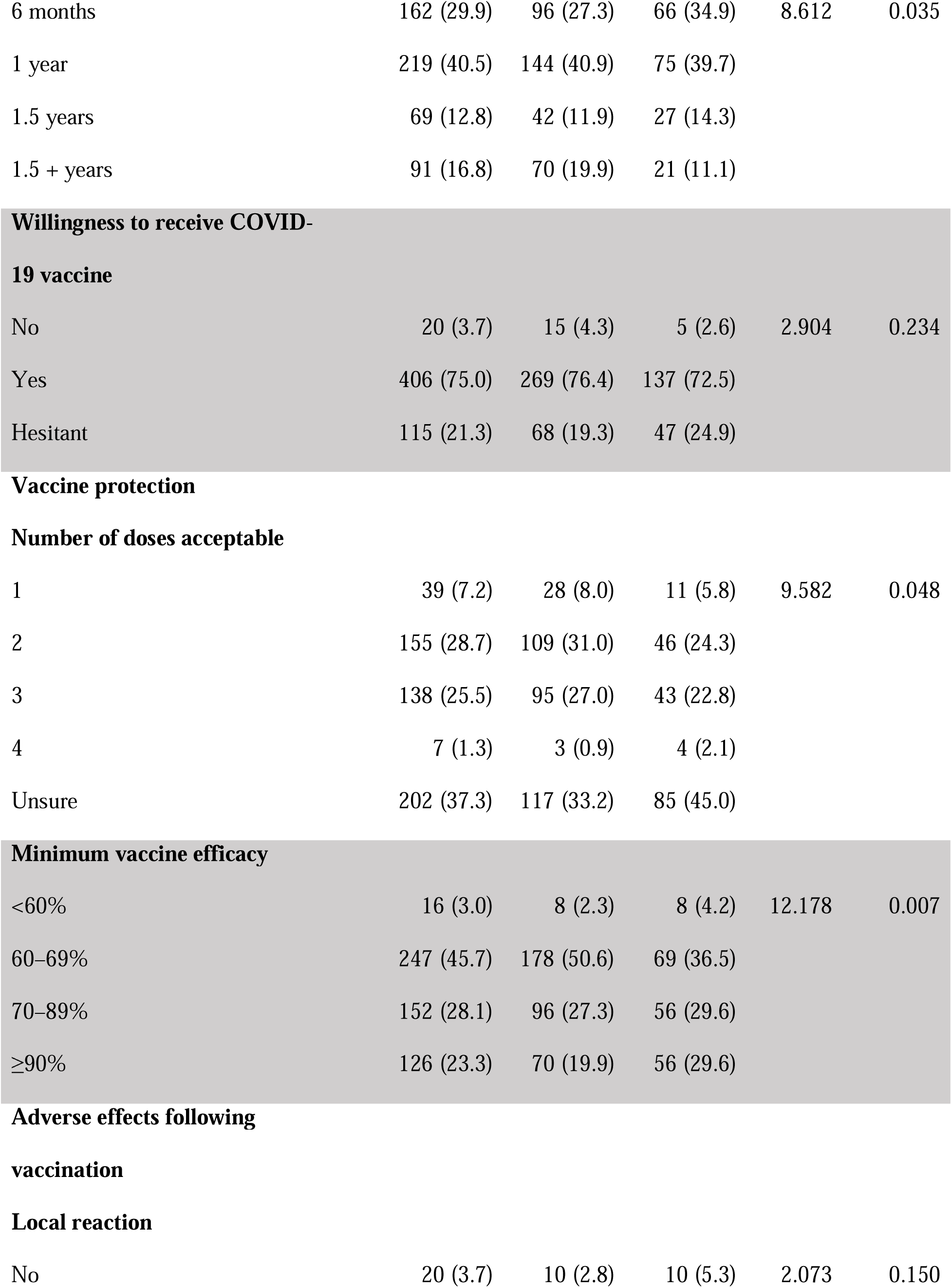
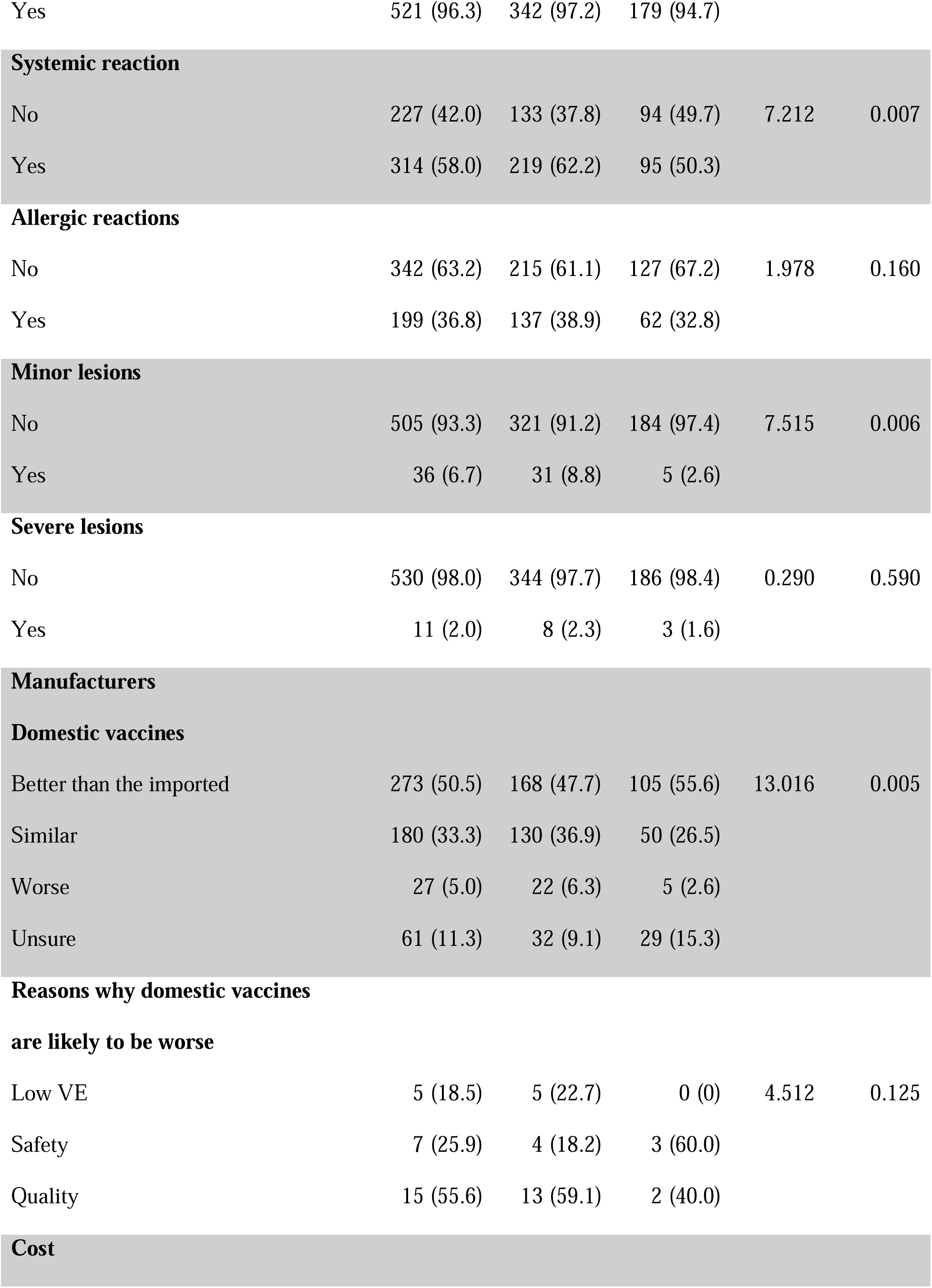
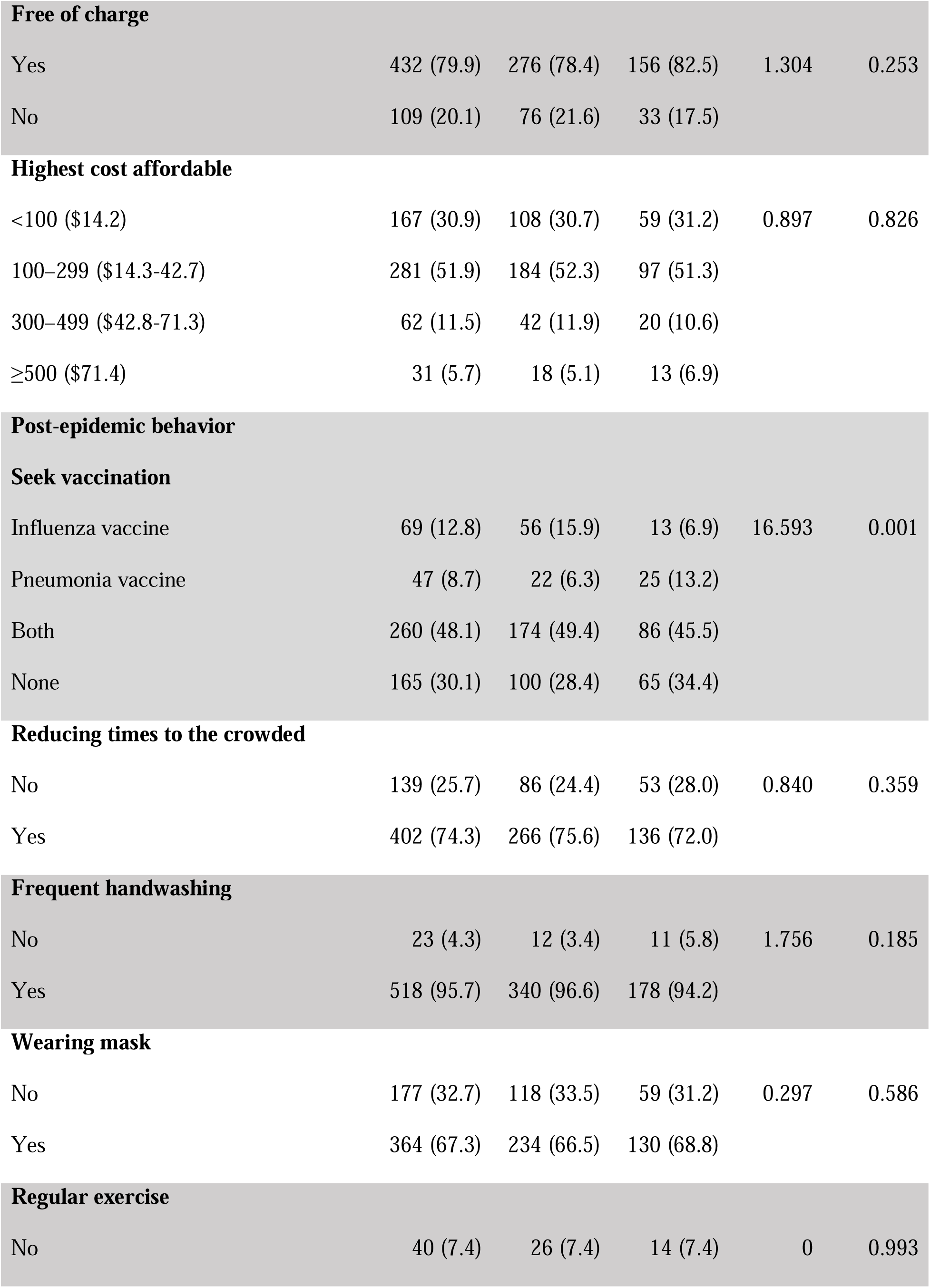
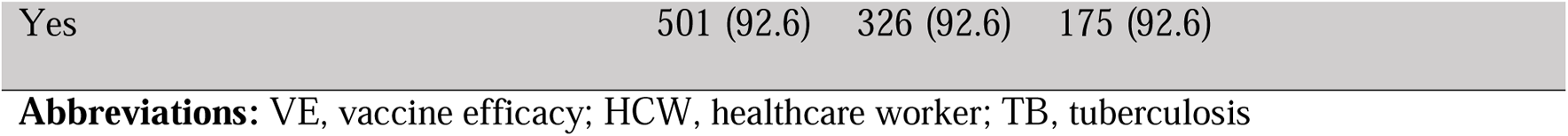
Characteristics of the participants and knowledge of disease, attitude, and acceptance of COVID-19 vaccination

| | Total | HCWs | General | $\chi^2$ | *p* |
| --- | --- | --- | --- | --- | --- |
| Demographics |  |  |  |  |  |
| Area |  |  |  |  |  |
| (i) high-level epidemic | 303 (56.0) | 170 (48.3) | 133 (70.4) | 24.321 | <0.001 |
| (ii)low-level epidemic | 238 (44.0) | 182 (51.7) | 56 (29.6) |  |  |
| Gender |  |  |  |  |  |
| Male | 230 (42.5) | 145 (41.2) | 85 (45.0) | 0.719 | 0.396 |
| Female | 311 (57.5) | 207 (58.8) | 104 (55.0) |  |  |
| Education |  |  |  |  |  |
| Masters or higher | 149 (27.5) | 114 (32.4) | 35 (18.5) | 65.057 | <0.001 |
| Bachelor | 276 (51.0) | 199 (56.5) | 77 (40.7) |  |  |
| Low than bachelor | 116 (21.4) | 39 (11.1) | 77 (40.7) |  |  |
| Age group (years) |  |  |  |  |  |
| 20–29 | 212 (39.2) | 130 (36.9) | 82 (43.4) | 4.834 | 0.184 |
| 30–39 | 175 (32.3) | 112 (31.8) | 63 (33.3) |  |  |
| 40–49 | 106 (19.6) | 78 (22.2) | 28 (14.8) |  |  |
| 50–59 | 48 (8.9) | 32 (9.1) | 16 (8.5) |  |  |
| Knowledge and attitude to disease |  |  |  |  |  |
| Vulnerable groups |  |  |  |  |  |
| All | 463 (85.6) | 305 (86.6) | 158 (83.6) | 0.927 | 0.336 |
| Subgroups | 78 (14.4) | 47 (13.4) | 31 (16.4) |  |  |
| Easily die after infection |  |  |  |  |  |
| Elderly or with chronic diseases | 537 (99.3) | 349 (99.1) | 188 (99.5) | 0.175 | 0.676 |
| Other | 4 (0.7) | 3 (0.9) | 1 (0.5) |  |  |
| **Effective drug for treatment** |  |  |  |  |  |
| No | 478 (88.4) | 325 (92.3) | 153 (81.0) | 28.789 | <0.001 |
| Yes | 20 (3.7) | 15 (4.3) | 5 (2.6) |  |  |
| Do not know | 43 (7.9) | 12 (3.4) | 31 (16.4) |  |  |
| **Virus mutation** |  |  |  |  |  |
| No | 32 (5.9) | 14 (4.0) | 18 (9.5) | 20.861 | <0.001 |
| Slight | 454 (83.9) | 314 (89.2) | 140 (74.1) |  |  |
| Dramatic | 55 (10.2) | 24 (6.8) | 31 (16.4) |  |  |
| **Possible trends of disease** |  |  |  |  |  |
| Diminishing | 98 (18.1) | 67 (19.0) | 31 (16.4) | 13.219 | 0.004 |
| Seasonal circulation | 79 (14.6) | 63 (17.9) | 16 (8.5) |  |  |
| Persistent epidemic | 24 (4.4) | 11 (3.1) | 13 (6.9) |  |  |
| Unsure | 340 (62.8) | 211 (59.9) | 129 (68.3) |  |  |
| **Possibility of future infection** |  |  |  |  |  |
| No | 210 (38.8) | 119 (33.8) | 91 (48.1) | 10.65 | 0.001 |
| Possible | 331 (61.2) | 233 (66.2) | 98 (51.9) |  |  |
| **COVID-19 vaccine** |  |  |  |  |  |
| **Necessity of vaccine development** |  |  |  |  |  |
| Yes | 517 (95.6) | 335 (95.2) | 182 (96.3) | 1.001 | 0.606 |
| No | 10 (1.8) | 8 (2.3) | 2 (1.1) |  |  |
| Not sure | 14 (2.6) | 9 (2.6) | 5 (2.6) |  |  |
| **Time to availability** |  |  |  |  |  |
| 6 months | 162 (29.9) | 96 (27.3) | 66 (34.9) | 8.612 | 0.035 |
| 1 year | 219 (40.5) | 144 (40.9) | 75 (39.7) |  |  |
| 1.5 years | 69 (12.8) | 42 (11.9) | 27 (14.3) |  |  |
| 1.5 + years | 91 (16.8) | 70 (19.9) | 21 (11.1) |  |  |

| **Willingness to receive COVID-19 vaccine** |  |  |  |  |  |
| --- | --- | --- | --- | --- | --- |
| No | 20 (3.7) | 15 (4.3) | 5 (2.6) | 2.904 | 0.234 |
| Yes | 406 (75.0) | 269 (76.4) | 137 (72.5) |  |  |
| Hesitant | 115 (21.3) | 68 (19.3) | 47 (24.9) |  |  |
| **Number of doses acceptable** |  |  |  |  |  |
| 1 | 39 (7.2) | 28 (8.0) | 11 (5.8) | 9.582 | 0.048 |
| 2 | 155 (28.7) | 109 (31.0) | 46 (24.3) |  |  |
| 3 | 138 (25.5) | 95 (27.0) | 43 (22.8) |  |  |
| 4 | 7 (1.3) | 3 (0.9) | 4 (2.1) |  |  |
| Unsure | 202 (37.3) | 117 (33.2) | 85 (45.0) |  |  |

| **Minimum vaccine efficacy** |  |  |  |  |  |
| --- | --- | --- | --- | --- | --- |
| <60% | 16 (3.0) | 8 (2.3) | 8 (4.2) | 12.178 | 0.007 |
| 60–69% | 247 (45.7) | 178 (50.6) | 69 (36.5) |  |  |
| 70–89% | 152 (28.1) | 96 (27.3) | 56 (29.6) |  |  |
| ≥90% | 126 (23.3) | 70 (19.9) | 56 (29.6) |  |  |
| **Local reaction** |  |  |  |  |  |
| No | 20 (3.7) | 10 (2.8) | 10 (5.3) | 2.073 | 0.150 |
| Yes | 521 (96.3) | 342 (97.2) | 179 (94.7) |  |  |
| **Systemic reaction** |  |  |  |  |  |
| No | 227 (42.0) | 133 (37.8) | 94 (49.7) | 7.212 | 0.007 |
| Yes | 314 (58.0) | 219 (62.2) | 95 (50.3) |  |  |
| **Allergic reactions** |  |  |  |  |  |
| No | 342 (63.2) | 215 (61.1) | 127 (67.2) | 1.978 | 0.160 |
| Yes | 199 (36.8) | 137 (38.9) | 62 (32.8) |  |  |
| **Minor lesions** |  |  |  |  |  |
| No | 505 (93.3) | 321 (91.2) | 184 (97.4) | 7.515 | 0.006 |
| Yes | 36 (6.7) | 31 (8.8) | 5 (2.6) |  |  |
| **Severe lesions** |  |  |  |  |  |
| No | 530 (98.0) | 344 (97.7) | 186 (98.4) | 0.290 | 0.590 |
| Yes | 11 (2.0) | 8 (2.3) | 3 (1.6) |  |  |
| **Manufacturers** |  |  |  |  |  |
| **Domestic vaccines** |  |  |  |  |  |
| Better than the imported | 273 (50.5) | 168 (47.7) | 105 (55.6) | 13.016 | 0.005 |
| Similar | 180 (33.3) | 130 (36.9) | 50 (26.5) |  |  |
| Worse | 27 (5.0) | 22 (6.3) | 5 (2.6) |  |  |
| Unsure | 61 (11.3) | 32 (9.1) | 29 (15.3) |  |  |
| **Reasons why domestic vaccines** |  |  |  |  |  |
| **are likely to be worse** |  |  |  |  |  |
| Low VE | 5 (18.5) | 5 (22.7) | 0 (0) | 4.512 | 0.125 |
| Safety | 7 (25.9) | 4 (18.2) | 3 (60.0) |  |  |
| Quality | 15 (55.6) | 13 (59.1) | 2 (40.0) |  |  |
| **Cost** |  |  |  |  |  |
| **Free of charge** |  |  |  |  |  |
| Yes | 432 (79.9) | 276 (78.4) | 156 (82.5) | 1.304 | 0.253 |
| No | 109 (20.1) | 76 (21.6) | 33 (17.5) |  |  |
| **Highest cost affordable** |  |  |  |  |  |
| <100 (\$14.2) | 167 (30.9) | 108 (30.7) | 59 (31.2) | 0.897 | 0.826 |
| 100–299 (\$14.3-42.7) | 281 (51.9) | 184 (52.3) | 97 (51.3) | | |
| 300–499 (\$42.8-71.3) | 62 (11.5) | 42 (11.9) | 20 (10.6) | | |
| ≥500 (\$71.4) | 31 (5.7) | 18 (5.1) | 13 (6.9) | | |
| **Post-epidemic behavior** |  |  |  |  |  |
| **Seek vaccination** |  |  |  |  |  |
| Influenza vaccine | 69 (12.8) | 56 (15.9) | 13 (6.9) | 16.593 | 0.001 |
| Pneumonia vaccine | 47 (8.7) | 22 (6.3) | 25 (13.2) |  |  |
| Both | 260 (48.1) | 174 (49.4) | 86 (45.5) |  |  |
| None | 165 (30.1) | 100 (28.4) | 65 (34.4) |  |  |
| **Reducing times to the crowded** |  |  |  |  |  |
| No | 139 (25.7) | 86 (24.4) | 53 (28.0) | 0.840 | 0.359 |
| Yes | 402 (74.3) | 266 (75.6) | 136 (72.0) |  |  |
| **Frequent handwashing** |  |  |  |  |  |
| No | 23 (4.3) | 12 (3.4) | 11 (5.8) | 1.756 | 0.185 |
| Yes | 518 (95.7) | 340 (96.6) | 178 (94.2) |  |  |
| **Wearing mask** |  |  |  |  |  |
| No | 177 (32.7) | 118 (33.5) | 59 (31.2) | 0.297 | 0.586 |
| Yes | 364 (67.3) | 234 (66.5) | 130 (68.8) |  |  |
| **Regular exercise** |  |  |  |  |  |
| No | 40 (7.4) | 26 (7.4) | 14 (7.4) | 0 | 0.993 |

|  |  |  |  |
| --- | --- | --- | --- |
| Yes | 501 (92.6) | 326 (92.6) | 175 (92.6) |

**Abbreviations:** VE, vaccine efficacy; HCW, healthcare worker; TB, tuberculosis

### 3.2 Knowledge of and attitude toward COVID-19

The majority of HCWs recognized that all age groups were susceptible to SARS-CoV-2 (n=305, 86.6%), and there was an increased risk of death in infected individuals who were older or had chronic diseases (n=349, 99.1%). Nearly 90% (314/352) of the HCWs believed that there would be slight mutations in the virus from the time it was first identified, and 7% (24/352) believed there were dramatic mutations. Of the 141 HCWs who reported their views about future COVID-19 trends, 47.5% (n=67) believed that incidence would diminish or disappear by summer (like SARS), 44.7% (n=63) believed that it would show annual cycles in incidence (like seasonal influenza), and only 8% (n=11) believed that it would become a endemic disease with year-round circulation (like tuberculosis). With regard to the risk of COVID-19, 66%(233/352) of the HCWs thought that they were at risk of becoming infected in the future, which was much higher than the equivalent proportion in the general population (52%, 98/189) (χ^2^=10.65, *p*<0.001).

### 3.3 Expectation about and acceptance of COVID-19 vaccination

With regard to the COVID-19 vaccines that were under development at the time of the survey, 95% of the HCWs thought that it was essential to be vaccinated. HCWs showed a higher acceptance to being vaccinated by a future vaccine. Compared to the general population, the HCWs believed that more time would be needed before vaccines were ready to be introduced for population-wide use (χ^2^=8.612, *p*=0.035), would accept a lower minimum threshold for vaccine effectiveness (χ^2^=12.178, *p*=0.007), more severe adverse effects (χ^2^=7.212, *p*=0.007), and more minor lesions (in the hypothetical situation that vaccination caused lesions) (χ^2^=7.515, *p*=0.006) than the general population (Table 2).

Approximately half the HCWs surveyed (47.7%, 168/352) believed that the COVID-19 vaccines produced by domestic manufacturers would be better than those produced abroad, which was lower than in the general population (55.6%, 105/189) (χ^2^=13.016, *p*=0.005). When asked about reasons for the potential inferiority of domestic vaccines among those who believed that imported vaccines would be better, approximately 60% (13/22) of HCWs suggested that this could be due to lower quality due to less stringently controlled production processes, and 60% (3/5) of the general population respondents believed that vaccine safety could not be fully guaranteed. In the entire study population, 78% (276/352) of HCWs agreed that the vaccine should be available free of charge although nearly half could afford to pay a price of 100–299 RMB (US$14–42) for a full course of vaccination. Three quarters (76.4%, 269/352) of the HCWs would opt to receive vaccination against COVID-19; however, nearly one fifth (19.3%, 68/352) were hesitant and needed more information before they could make a decision.

### 3.4 Protection adopted post epidemic

Approximately 70% (252/352) of the HCWs intended to be vaccinated against influenza and/or *Streptococcus pneumoniae*. The HCWs preferred to receive influenza vaccination (65.3%, 230/352), whereas the general population preferred to receive pneumococcal vaccination (58.7%, 111/189) (*p*=0.001). Both HCWs and the general population indicated that they would decrease the frequency of venturing into crowed areas (74%, 402/541), wash their hands more frequently (96%, 518/541), participate in physical exercise (93%, 501/541), and wear a face covering (67%, 364/541) in the future.

### 3.5 Vaccine preferences

As shown in Table 3, the DCE respondents indicated that the largest preference for a decision to receive COVID-19 vaccination (*all* p<0.001) are a seasonal (OR 4.37, 95% confidence interval [CI] 3.72–5.13) or persistent (OR 3.07, 95% CI 2.61–3.61) COVID-19 epidemic; a high vaccine refusal rate among social contacts of the respondents (OR 0.40, 95% CI 0.34– 0.47), neutrality (OR 0.41, 95% CI 0.35–0.49), high risk of infection (>30%; OR: 2.08, 95% CI 1.78–2.43), fee of 300 RMB (OR 0.48, 95% CI 0.40–0.56) and 100 RMB (OR 0.58, 95% CI 0.50–0.67); vaccine efficacy >80% (OR 1.77, 95% CI 1.54–2.04) and mild side effects only following immunisation (OR 1.54, 95% CI 1.35–1.75).

**Table 3.**
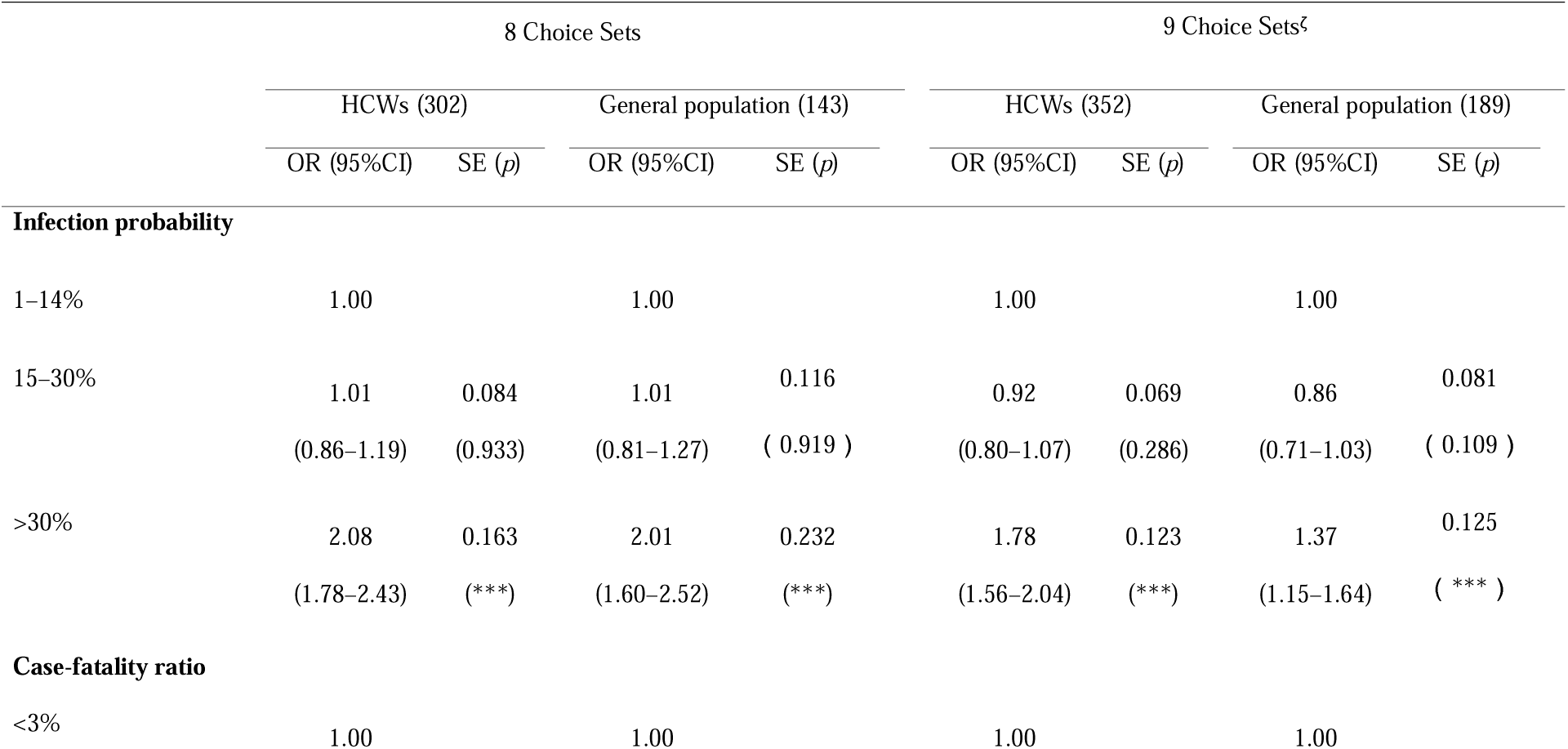
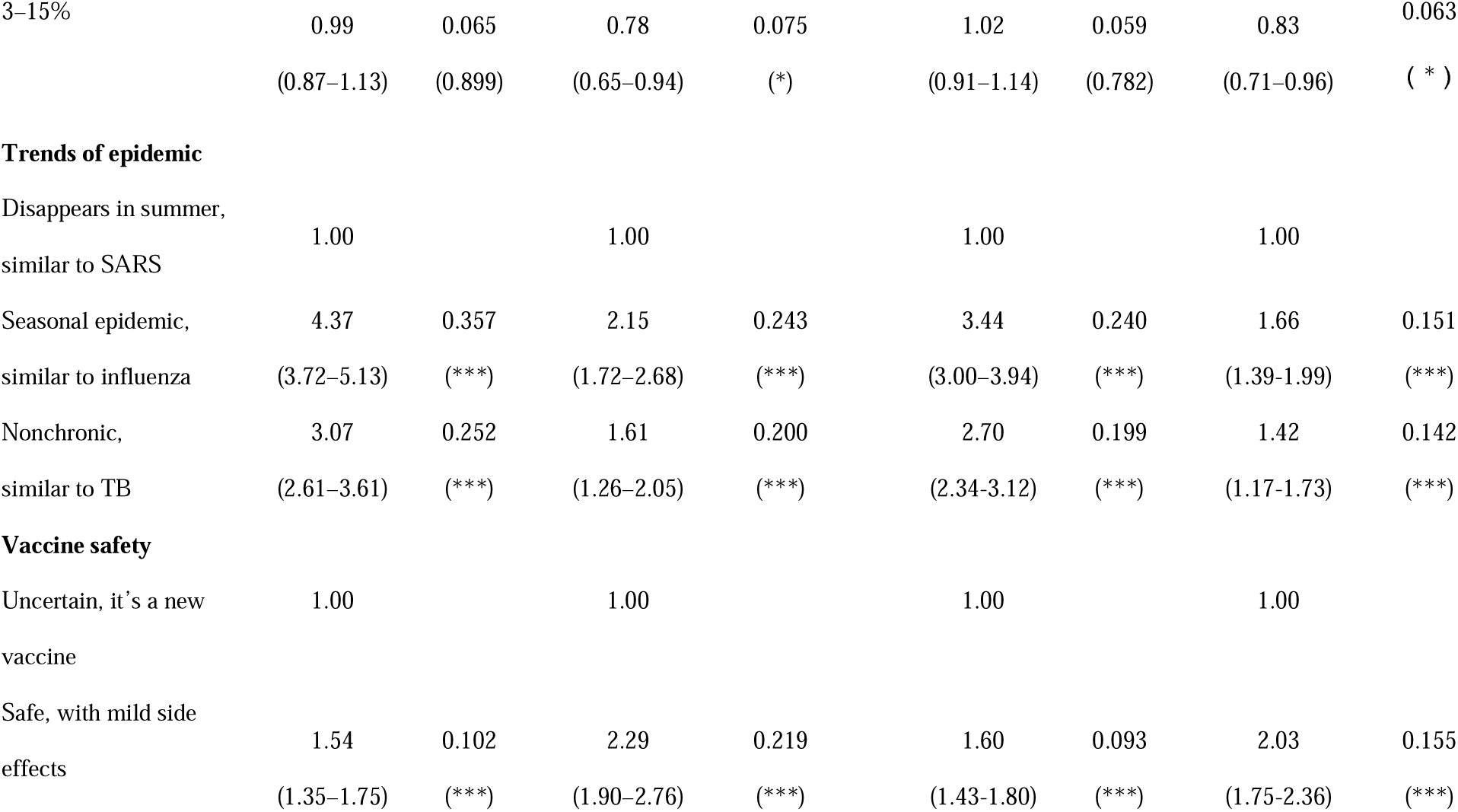
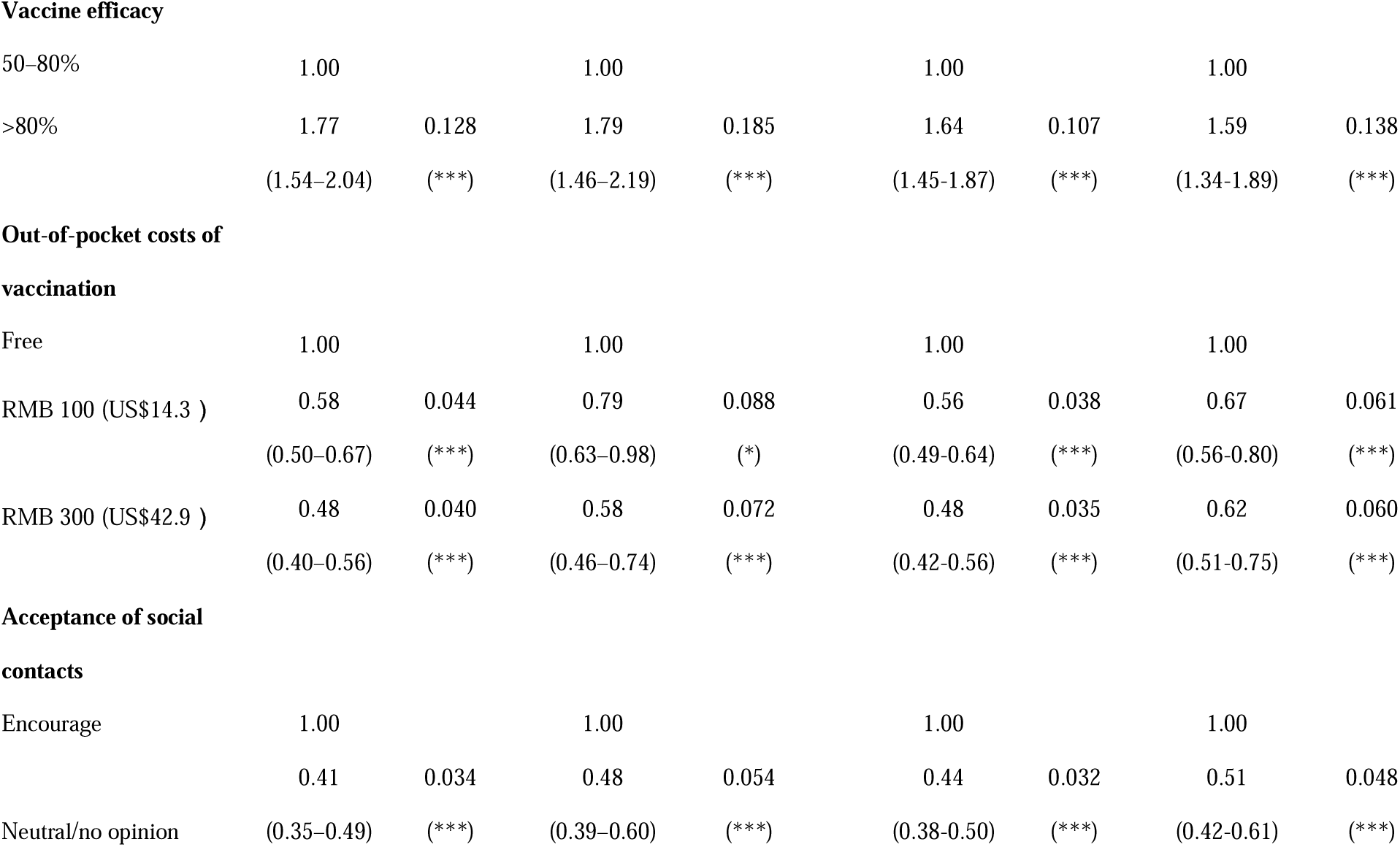
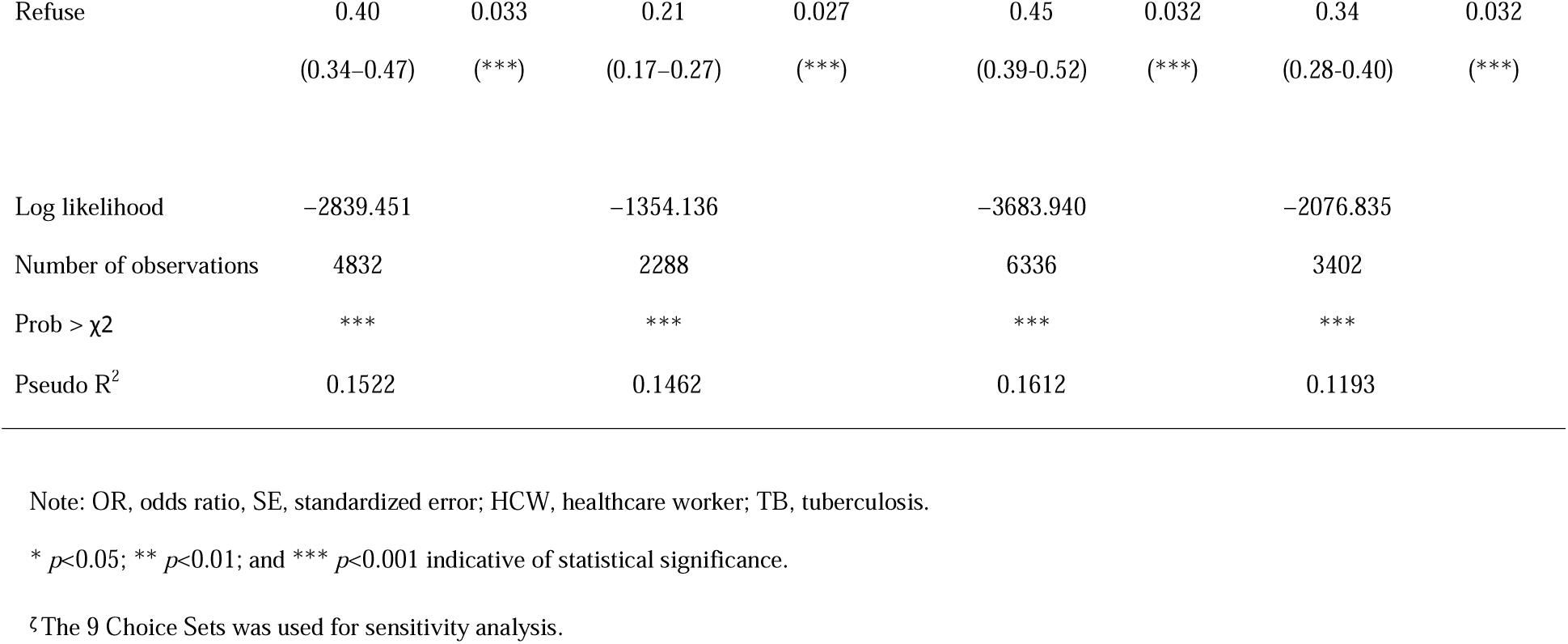
Preferences for COVID-19 vaccination among HCWs and the general population

|  | 8 Choice Sets |  |  |  | 9 Choice Sets<sup>‡</sup> |  |  |  |
| --- | --- | --- | --- | --- | --- | --- | --- | --- |
|  | HCWs (302) |  | General population (143) |  | HCWs (352) |  | General population (189) |  |
|  | OR (95%CI) | SE (*p*) | OR (95%CI) | SE (*p*) | OR (95%CI) | SE (*p*) | OR (95%CI) | SE (*p*) |
| **Infection probability** |  |  |  |  |  |  |  |  |
| 1–14% | 1.00 |  | 1.00 |  | 1.00 |  | 1.00 |  |
| 15–30% | 1.01 | 0.084 | 1.01 | 0.116 | 0.92 | 0.069 | 0.86 | 0.081 |
|  | (0.86–1.19) | (0.933) | (0.81–1.27) | ( 0.919 ) | (0.80–1.07) | (0.286) | (0.71–1.03) | ( 0.109 ) |
| >30% | 2.08 | 0.163 | 2.01 | 0.232 | 1.78 | 0.123 | 1.37 | 0.125 |
|  | (1.78–2.43) | (***) | (1.60–2.52) | (***) | (1.56–2.04) | (***) | (1.15–1.64) | ( ***) ) |
| **Case-fatality ratio** |  |  |  |  |  |  |  |  |
| <3% | 1.00 |  | 1.00 |  | 1.00 |  | 1.00 |  |
| 3–15% | 0.99 | 0.065 | 0.78 | 0.075 | 1.02 | 0.059 | 0.83 | 0.063 |
|  | (0.87–1.13) | (0.899) | (0.65–0.94) | (*) | (0.91–1.14) | (0.782) | (0.71–0.96) | (*) |
| **Trends of epidemic** |  |  |  |  |  |  |  |  |
| Disappears in summer,<br>similar to SARS | 1.00 |  | 1.00 |  | 1.00 |  | 1.00 |  |
| Seasonal epidemic,<br>similar to influenza | 4.37 | 0.357 | 2.15 | 0.243 | 3.44 | 0.240 | 1.66 | 0.151 |
|  | (3.72–5.13) | (***) | (1.72–2.68) | (***) | (3.00–3.94) | (***) | (1.39-1.99) | (***) |
| Nonchronic,<br>similar to TB | 3.07 | 0.252 | 1.61 | 0.200 | 2.70 | 0.199 | 1.42 | 0.142 |
|  | (2.61–3.61) | (***) | (1.26–2.05) | (***) | (2.34-3.12) | (***) | (1.17-1.73) | (***) |
| **Vaccine safety** |  |  |  |  |  |  |  |  |
| Uncertain, it's a new<br>vaccine | 1.00 |  | 1.00 |  | 1.00 |  | 1.00 |  |
| Safe, with mild side<br>effects | 1.54 | 0.102 | 2.29 | 0.219 | 1.60 | 0.093 | 2.03 | 0.155 |
|  | (1.35–1.75) | (***) | (1.90–2.76) | (***) | (1.43-1.80) | (***) | (1.75-2.36) | (***) |
| **Vaccine efficacy** |  |  |  |  |  |  |  |  |
| 50–80% | 1.00 |  | 1.00 |  | 1.00 |  | 1.00 |  |
| >80% | 1.77 | 0.128 | 1.79 | 0.185 | 1.64 | 0.107 | 1.59 | 0.138 |
|  | (1.54–2.04) | (***) | (1.46–2.19) | (***) | (1.45-1.87) | (***) | (1.34-1.89) | (***) |
| **Out-of-pocket costs of vaccination** |  |  |  |  |  |  |  |  |
| Free | 1.00 |  | 1.00 |  | 1.00 |  | 1.00 |  |
| RMB 100 (US\$14.3 ) | 0.58 | 0.044 | 0.79 | 0.088 | 0.56 | 0.038 | 0.67 | 0.061 |
|  | (0.50–0.67) | (***) | (0.63–0.98) | (*) | (0.49-0.64) | (***) | (0.56-0.80) | (***) |
| RMB 300 (US\$42.9 ) | 0.48 | 0.040 | 0.58 | 0.072 | 0.48 | 0.035 | 0.62 | 0.060 |
|  | (0.40–0.56) | (***) | (0.46–0.74) | (***) | (0.42-0.56) | (***) | (0.51-0.75) | (***) |
| **Acceptance of social contacts** |  |  |  |  |  |  |  |  |
| Encourage | 1.00 |  | 1.00 |  | 1.00 |  | 1.00 |  |
|  | 0.41 | 0.034 | 0.48 | 0.054 | 0.44 | 0.032 | 0.51 | 0.048 |
| Neutral/no opinion | (0.35–0.49) | (***) | (0.39–0.60) | (***) | (0.38-0.50) | (***) | (0.42-0.61) | (***) |
| Refuse | 0.40<br>(0.34–0.47) | 0.033<br>(***) | 0.21<br>(0.17–0.27) | 0.027<br>(***) | 0.45<br>(0.39–0.52) | 0.032<br>(***) | 0.34<br>(0.28–0.40) | 0.032<br>(***) |
| Log likelihood | –2839.451 |  | –1354.136 |  | –3683.940 |  | –2076.835 |  |
| Number of observations | 4832 |  | 2288 |  | 6336 |  | 3402 |  |
| Prob > $\chi^2$ | *** | | *** | | *** | | *** | |
| Pseudo R<sup>2</sup> | 0.1522 |  | 0.1462 |  | 0.1612 |  | 0.1193 |  |

Note: OR, odds ratio, SE, standardized error; HCW, healthcare worker; TB, tuberculosis.
\* $p < 0.05$ ; \*\* $p < 0.01$ ; and \*\*\* $p < 0.001$ indicative of statistical significance.
‡ The 9 Choice Sets was used for sensitivity analysis.

The decision to get vaccinated were reported in the general population, although the odds ratios were generally smaller. However, the general population respondents focus more on vaccine safety (OR 2.29, *p*<0.001), decision to vaccinate among social contacts (OR 0.21 and 0.48, *p*<0.001), and the case-fatality ratio (OR 0.78, *p*<0.05) than the HCWs. A sensitivity analysis in which all participants were included regardless of whether they answered the consistency question correctly, gave effects for both HCWs and the general population (see Table 3).

## 4. Discussion

This is one of the first studies to measure the acceptance of and preference for COVID-19 vaccination in HCWs. Using an internet-based design, we surveyed the HCW’s knowledge of and attitudes toward COVID-19 and their acceptance of a future SARS-CoV-2 vaccine from 26 provinces in China. Compared to the general population, vaccination decisions indicated by the HCWs were less sensitive to adverse effects and vaccine efficacy. The experience of the COVID-19 pandemic and related health education can decrease vaccine hesitancy for other vaccine preventable diseases such as influenza and pneumonia. Future disease trends, vaccination decisions of social contacts, and the risk of becoming infected were more important drivers of choosing vaccination among HCWs.

### 4.1 HCWs’ attitude to and willingness for COVID-19 vaccination

A similar percentage of HCW and the general population respondents (76.4% and 72.5%) were willing to get vaccinated. It is believed that the Chinese have much higher confidence in vaccination, especially for the Expanded Immunization (EPI) vaccines, than many other parts of the world. Although the COVID-19 vaccine was not included in the EPI list till now, the public in China are prone to have less hesitancy and more uptake. For the general population, China had much higher COVID-19 vaccine acceptance (91%) compared to most countries (range 38-98%)^20^. According to a survey conducted in May 2020, 49% of US respondents reported willingness to get vaccinated against COVID-19, which is much lower than the proportion in this study. Furthermore, 31% of US respondents were vaccine hesitant, a higher proportion than in this study (19.3% of HCWs and 24.9% of the general population) ^21^. The proportion of respondents in our study willing the get vaccinated was also higher than reported in some other countries during the 2009 H1N1 pandemic (56.1% in the UK, 64% in the US, and 54.7% in Australia)^19, 22, 23^.

COVID-19 vaccines were approved in late 2020 and early 2021 for population use in many countries across the world. During that time, HCWs’ attitudes towards COVID-19 vaccination has varied across countries. HCWs’ hesitancy toward COVID-19 may also vary over time. Overall Chinese HCWs seems to have more confidence and willingness to receive the vaccine than many parts of the world. A survey conducted in France, French-speaking Belgium and Quebec, Canada reported 71.6% acceptance of COVID-19 vaccination among HCWs^24^. A study in October and November 2020 showed that 36% of US HCWs were willing to receive COVID-19 vaccine as soon as it became available, while 56% were hesitant^25^. In Germany, 57% of front-line HCWs in emergency medical services (EMS) were willing to be vaccinated in a survey from December 2020 to January 2021^26^. A systematic review indicated that 22.5% out of 76 471 HCWs worldwide reported COVID-19 vaccine hesitancy, with the proportion ranging between countries from 4.3% to 72%. Concerns about vaccine safety, efficacy and potential adverse effects were the leading reasons for hesitancy in the survey^27^. Hence, continued surveillance of the attitudes of HCWs from diverse socioeconomic and geographical backgrounds is essential.

Most HCWs expressed willingness to receive COVID-19 vaccination after vaccine introduction, which, to a large extent, is attributable to their risk perception of infection and confidence in the efficacy and safety of the vaccine. HCWs have a much higher risk of getting infected with SARS-CoV-2 compared to the general population. For example, by Feb 11, 2020, 6 deaths and 1716 infections in HCWs occurred in mainland China (0.4% and 3.8% of the total deaths and infection respectively) ^28^. In this study, 66% of the HCWs believed that they were at risk of being infected by SARS-CoV-2 from close contact with COVID-19 patients. Furthermore, uncertainty around how the epidemic may develop contributed to the risk evaluation: 60% of HCWs in the study were uncertain about the future trend in COVID-19 epidemics.

The HCWs showed a positive attitude toward the vaccine and were willing to accept a vaccine with effectiveness as low as 60–70% (similar to seasonal influenza vaccines), more severe adverse effects, and a higher number of doses than respondents from the general population. HCWs are vital to the public’s vaccination decisions, so their attitudes to vaccination can influence overall vaccine coverage. Adequate knowledge about a vaccine has been found in previous studies to increase HCW’s willingness to recommend that vaccine. A study in the UK reported that nurses with high levels of knowledge about seasonal influenza vaccination were more likely to recommend both influenza vaccination and vaccination in general to their patients ^29^. In a study on HPV vaccination in Cameroon, one of the most important determinants of whether nurses would recommend the vaccine to the patients was their understanding of the efficacy and safety of the vaccine ^30^.

In this study, nearly 20% and 25% of HCWs and the general population surveyed, respectively, were hesitant about COVID-19 vaccination, which may hamper establishment of widespread vaccine-induced protection in the population. Lack of preparedness for advising patients about vaccination and lack of training can inhibit HCWs in making effective vaccine recommendations. Since HCWs play a key role in reducing the disease burden of COVID-19 and helping to increase vaccine coverage, efforts need to be invested in increasing their knowledge and acceptance of vaccination.

### 4.2 Preference and relevant factors for COVID-19 vaccination

We found that respondents’ expectation about the future trend in COVID-19 was far more influential on their decision to want to be vaccinated than their infection risk or the case-fatality ratio of COVID-19. Their preferences may be motivated by their experience of seasonal influenza vaccination, since the seasonal influenza virus displays annual seasonality in temperate regions. Participants in the study were also influenced to the safety of future COVID-19 vaccines.

The behavior and attitudes of social contacts such as relatives, friends, and neighbors, play an important role in the decision for both HCWs and the general population. When vaccine safety or efficacy is uncertain, external cues such as vaccination uptake by others may be highly influential on vaccination intent ^17^. Vaccine hesitancy has been one of the most important public health issues in many countries, including some of those in Europe and North America. In contrast, vaccine hesitancy has remained limited in China, and vaccine confidence has recovered rapidly after vaccine safety or quality-related incidents in the past^31, 32^.The reason for the resilience of vaccine confidence has been attributed by other authors to the collective culture in China where nonconformity is more strongly associated with deviance ^33^. This suggests that appealing to a communal responsibility to protect others via indirect protection may be a strategy to maintain COVID-19 vaccine uptake.

Surprisingly, disease severity was not found to have a strong relationship with vaccination decisions among HCWs, and actually showed a negative (although non-significant, with CIs for the OR overlapping 1.0) association in the general population sample. However, similar relationships have been observed with other vaccine-preventable diseases such as seasonal influenza ^17^.

Efficacy was found to be the factor that most influenced vaccine selection in a DCE survey in UK, especially for those aged ≥ 55 years^34^. An online DCE survey administered in China found that public prefer high effectiveness of the vaccine, followed by long protective duration, very few adverse events and being manufactured overseas and price was the least important attribute^35^. The studies support our findings that future COVID-19 disease incidence and infection risk could significantly increase the probability of vaccination acceptance in HCWs.

### 4.3 Attitude toward domestically produced COVID-19 vaccine

Due to concerns about the quality of domestically produced vaccines, some HCWs may be hesitant to be vaccinated, which could have knock on effects on the vaccine recommendations they make to their patients. In this study, the proportion of HCWs who considered domestic vaccines to be inferior was twice that in the general population (7% vs 3%). Sixty percent of Chinese HCWs were concerned about less stringent quality control of vaccine production rather than the vaccine itself. Vaccine quality incidents reported in China over the past decade may have contributed to a decline in confidence in domestically-produced vaccines among the Chinese population, including HCWs ^31, 36^. For example, on July 15, 2018, after an unannounced inspection of the Changchun Changsheng Biotech drug producer, the China Food and Drug Administration (CFDA) confirmed that fraudulent records were made during rabies vaccine production. Six days later, an article entitled “King of Vaccine” by “Beast Father” was posted in the WeChat media social account that received more than 2 million views within an hour of its release and triggered a public outcry. Rumors of ‘poisonous’ vaccines were quickly spread throughout online media ^36^. Like human immune memory which is reactivated following another challenge with a similar antigen, HCWs and the general public may react negatively toward a future COVID-19 vaccine if serious adverse events (e.g. allergic reactions and blood clots following vaccination seen recently) are reported after vaccine introduction.

### 4.4 Potential positive influence including seasonal influenza and pneumonia vaccination following the pandemic

The COVID-19 epidemic could be used as an opportunity to improve public awareness of and knowledge about other infectious disease threats. Study respondents indicated increased use of personal protective measures that could reduce the incidence of other infections, such as face covering use and uptake of non-COVID-19 vaccines. Public awareness of seasonal influenza burden has increased in China in recent years, as seen in the shortages of vaccine supplies experienced in recent years ^37, 38^. This awareness may have been heightened by the perception that SARS-CoV-2 was an “enhanced influenza” virus, and may have contributed to the increased willingness to receive seasonal influenza vaccination among HCWs. In the past, seasonal influenza vaccine coverage among Chinese HCWs has been low (17.7%, 95%CI 12.8–22.6) ^39^, so one positive development from the COVID-19 pandemic may be increased vaccine uptake for seasonal influenza. Among general population respondents, we saw higher uptake of pneumococcal vaccination, possibly because of fear that COVID-19 may seriously damage the lungs ^40^. In China, knowledge about the pneumococcal vaccine is poor and the coverage is very low. For example, in a survey conducted in the city of Hangzhou in 2013, only 21.8% of older adults were willing to receive the 23-valent pneumococcal polysaccharide vaccine (PPV23) vaccination, and actual coverage among older adults was only 1.23%^41^.

### 4.5 Limitations

Several limitations exist in the present study. First, the small sample was not randomly recruited and may not be representative of HCWs in the country; thus, any generalization of these results should be undertaken with caution. In particular, online recruitment via WeChat may have biased the study towards a better-educated and/or higher income sample. For example, only 36.2% of the HCWs in China have a bachelor’s degree or higher qualification, which is much lower than that in the present study (88.9%) ^42^. However, the HCW and general population samples are at least likely to suffer from the same kind of biases, since they were recruited simultaneously using the same methods, and have similar demographics in terms of gender and age distribution. The sample size is not large enough for us to report detailed interaction between attribute (levels) and individual characteristics. Second, participants were recruited and surveyed online instead of in a face-to-face interview, which may have led to potential biases. Third, we did not distinguish between different types of HCWs, such as doctors and nurses in hospitals, health care providers in the community, and staff in local Centers for Disease Control and Preventions, all of whom may have had different levels of knowledge about vaccination and decision-making influences. Fourth, the data could not be well run through the mixed logit model which we had designed for DCE. One possible reason is in that we lack relative data to determine quantitative variables in March 2020, which may hinder the inference and implication. Fifth, as a tool to measure the preferences, the DCE cannot include too many attributes and levels which makes it different from real situation. In addition, the consistency between the predicted results of the model and the actual selection results of individuals has not been confirmed.

## 5. Conclusions

Factors that contribute to COVID-19 vaccination decisions include personal risk perception, attitude towards vaccination in general, information sources, access and demographic variables, as well as practical factors ^7^. For COVID-19 vaccination programs in China and elsewhere need to address these factors in both HCWs and in the general population. Multi-component interventions to address the key determinants of demand and hesitancy should be considered. Education of HCWs should be prioritized since increased vaccine acceptance in this group may benefit both HCWs and the people they recommend vaccination to.

## Supporting information

eSupplements

## Data Availability

The datasets used and/or analyzed during the current study are available from the corresponding author on reasonable request.

## Authors’ contributions

CF designed and conducted the survey. ZW, YW, and XS collected data. CF, ZW and FZ wrote the manuscript. SP, SL, and PL conducted the analysis and revised the manuscript. MJ was as major contributor in writing the manuscript.

## Acknowledgements

Not applicable.

## References

1. WHO Coronavirus (COVID-19) Dashboard. Accessed June 9, 2021. https://covid19.who.int/

2. Li Q, Guan X, Wu P, et al. Early Transmission Dynamics in Wuhan, China, of Novel Coronavirus-Infected Pneumonia. N Engl J Med. Mar 26 2020;382(13):1199–1207. doi:10.1056/NEJMoa2001316

3. Li R, Pei S, Chen B, et al. Substantial undocumented infection facilitates the rapid dissemination of novel coronavirus (SARS-CoV2). Science. Mar 16 2020;doi:10.1126/science.abb3221

4. World Health Organization. COVID-19 vaccine tracker and landscape. Accessed June 8, 2021. https://www.who.int/publications/m/item/draft-landscape-of-covid-19-candidate-vaccines

5. World Health Organization. Improving vaccination demand and addressing hesitancy. Accessed August 16, 2019. https://www.who.int/immunization/programmes_systems/vaccine_hesitancy/en/

6. Larson HJ, Clarke RM, Jarrett C, et al. Measuring trust in vaccination: A systematic review. Hum Vaccin Immunother. Jul 3 2018;14(7):1599–1609. doi:10.1080/21645515.2018.1459252

7. WHO. Improving vaccination demand and addressing hesitancy. Updated 16 August 2019. Accessed 16 August, 2019. https://www.who.int/immunization/programmes_systems/vaccine_hesitancy/en/

8. World Health Organization. WHO SAGE ROADMAP FOR PRIORITIZING USES OF COVID-19 VACCINES IN THE CONTEXT OF LIMITED SUPPLY An approach to inform planning and subsequent recommendations based upon epidemiologic setting and vaccine supply scenarios. Accessed November 13, 2020. https://www.who.int/docs/default-source/immunization/sage/covid/sage-prioritization-roadmap-covid19-vaccines.pdf

9. COVID-19: protecting health-care workers. Lancet. Mar 21 2020;395(10228):922. doi:10.1016/S0140-6736(20)30644-9

10. Carman WF, Elder AG, Wallace LA, et al. Effects of influenza vaccination of health-care workers on mortality of elderly people in long-term care: a randomised controlled trial. Lancet. Jan 8 2000;355(9198):93–7. doi:10.1016/S0140-6736(99)05190-9

11. Dempsey AF, Pyrznawoski J, Lockhart S, et al. Effect of a Health Care Professional Communication Training Intervention on Adolescent Human Papillomavirus Vaccination: A Cluster Randomized Clinical Trial. JAMA Pediatr. May 7 2018;172(5):e180016. doi:10.1001/jamapediatrics.2018.0016

12. Kempe A, O’Leary ST, Markowitz LE, et al. HPV Vaccine Delivery Practices by Primary Care Physicians. Pediatrics. Oct 2019;144(4)doi:10.1542/peds.2019-1475

13. Paterson P, Meurice F, Stanberry LR, Glismann S, Rosenthal SL, Larson HJ. Vaccine hesitancy and healthcare providers. Vaccine. Dec 20 2016;34(52):6700–6706. doi:10.1016/j.vaccine.2016.10.042

14. de Bekker-Grob EW, Ryan M, Gerard K. Discrete choice experiments in health economics: a review of the literature. Health Econ. Feb 2012;21(2):145–72. doi:10.1002/hec.1697

15. Brown DS, Poulos C, Johnson FR, Chamiec-Case L, Messonnier ML. Adolescent girls’ preferences for HPV vaccines: a discrete choice experiment. Adv Health Econ Health Serv Res. 2014;24:93–121.

16. Eilers R, de Melker HE, Veldwijk J, Krabbe PFM. Vaccine preferences and acceptance of older adults. Vaccine. May 15 2017;35(21):2823–2830. doi:10.1016/j.vaccine.2017.04.014

17. Liao Q, Lam WWT, Wong CKH, Lam C, Chen J, Fielding R. The relative effects of determinants on Chinese adults’ decision for influenza vaccination choice: What is the effect of priming? Vaccine. Jul 9 2019;37(30):4124–4132. doi:10.1016/j.vaccine.2019.05.072

18. National Health Committee of the People’s Republic of China. Update on COVID-19 as of 24:00 on Mar 15. http://www.nhc.gov.cn/xcs/yqtb/202003/114113d25c1d47aabe68381e836f06a8.shtml

19. Horney JA, Moore Z, Davis M, MacDonald PD. Intent to receive pandemic influenza A (H1N1) vaccine, compliance with social distancing and sources of information in NC, 2009. PLoS One. Jun 18 2010;5(6):e11226. doi:10.1371/journal.pone.0011226

20. Wouters OJ, Shadlen KC, Salcher-Konrad M, et al. Challenges in ensuring global access to COVID-19 vaccines: production, affordability, allocation, and deployment. Lancet. Mar 13 2021;397(10278):1023–1034. doi:10.1016/S0140-6736(21)00306-8

21. The Associated Press-NORC Center for Public Affairs Research. Expectations for a COVID-19 Vaccine. Accessed May 28, 2020. http://www.apnorc.org/projects/Pages/Expectations-for-a-COVID-19-Vaccine.aspx

22. Nguyen T, Henningsen KH, Brehaut JC, Hoe E, Wilson K. Acceptance of a pandemic influenza vaccine: a systematic review of surveys of the general public. Infect Drug Resist. 2011;4:197–207. doi:10.2147/IDR.S23174

23. Rubin GJ, Potts HW, Michie S. The impact of communications about swine flu (influenza A H1N1v) on public responses to the outbreak: results from 36 national telephone surveys in the UK. Health Technol Assess. Jul 2010;14(34):183–266. doi:10.3310/hta14340-03

24. Verger P, Scronias D, Dauby N, et al. Attitudes of healthcare workers towards COVID-19 vaccination: a survey in France and French-speaking parts of Belgium and Canada, 2020. Euro Surveill. Jan 2021;26(3)doi:10.2807/1560-7917.ES.2021.26.3.2002047

25. Shekhar R, Sheikh AB, Upadhyay S, et al. COVID-19 Vaccine Acceptance among Health Care Workers in the United States. Vaccines (Basel). Feb 3 2021;9(2)doi:10.3390/vaccines9020119

26. Nohl A, Afflerbach C, Lurz C, et al. Acceptance of COVID-19 Vaccination among Front-Line Health Care Workers: A Nationwide Survey of Emergency Medical Services Personnel from Germany. Vaccines (Basel). Apr 23 2021;9(5)doi:10.3390/vaccines9050424

27. Biswas N, Mustapha T, Khubchandani J, Price JH. The Nature and Extent of COVID-19 Vaccination Hesitancy in Healthcare Workers. J Community Health. Apr 20 2021;doi:10.1007/s10900-021-00984-3

28. National Health Committee of the People’s Republic of China. State Council press conference on Feb 14 2020. http://www.nhc.gov.cn/xwzb/webcontroller.do?titleSeq=11231&gecstype=1

29. Zhang J, While AE, Norman IJ. Nurses’ vaccination against pandemic H1N1 influenza and their knowledge and other factors. Vaccine. Jul 6 2012;30(32):4813–9. doi:10.1016/j.vaccine.2012.05.012

30. Wamai RG, Ayissi CA, Oduwo GO, et al. Awareness, knowledge and beliefs about HPV, cervical cancer and HPV vaccines among nurses in Cameroon: an exploratory study. Int J Nurs Stud. Oct 2013;50(10):1399–406. doi:10.1016/j.ijnurstu.2012.12.020

31. Yu W, Liu D, Zheng J, et al. Loss of confidence in vaccines following media reports of infant deaths after hepatitis B vaccination in China. Int J Epidemiol. Apr 2016;45(2):441–9. doi:10.1093/ije/dyv349

32. Tu S, Sun FY, Chantler T, et al. Caregiver and service provider vaccine confidence following the Changchun Changsheng vaccine incident in China: A cross-sectional mixed methods study. Vaccine. Oct 14 2020;38(44):6882–6888. doi:10.1016/j.vaccine.2020.08.063

33. Yang Y. Health research: social and behavioral theory and methods. 2nd ed. People Medical Publishing House; 2017.

34. McPhedran R, Toombs B. Efficacy or delivery? An online Discrete Choice Experiment to explore preferences for COVID-19 vaccines in the UK. Econ Lett. Mar 2021;200:109747. doi:10.1016/j.econlet.2021.109747

35. Dong D, Xu RH, Wong EL, et al. Public preference for COVID-19 vaccines in China: A discrete choice experiment. Health Expect. Dec 2020;23(6):1543–1578. doi:10.1111/hex.13140

36. Fu C. Milestone and challenges: lessons from defective vaccine incidents in China. Hum Vaccin Immunother. 2020;16(1):80. doi:10.1080/21645515.2019.1646579

37. Li L, Liu Y, Wu P, et al. Influenza-associated excess respiratory mortality in China, 2010-15: a population-based study. Lancet Public Health. Sep 2019;4(9):e473–e481. doi:10.1016/S2468-2667(19)30163-X

38. Ren X, Geoffroy E, Tian K, et al. Knowledge, Attitudes, and Behaviors (KAB) of Influenza Vaccination in China: A Cross-Sectional Study in 2017/2018. Vaccines (Basel). Dec 26 2019;8(1)doi:10.3390/vaccines8010007

39. Wang Q, Yue N, Zheng M, et al. Influenza vaccination coverage of population and the factors influencing influenza vaccination in mainland China: A meta-analysis. Vaccine. Nov 19 2018;36(48):7262–7269. doi:10.1016/j.vaccine.2018.10.045

40. Zhan S, Yang YY, Fu C. Public’s early response to the novel coronavirus-infected pneumonia. Emerg Microbes Infect. 2020;9(1):534. doi:10.1080/22221751.2020.1732232

41. Liu S, Xu E, Liu Y, et al. Factors associated with pneumococcal vaccination among an urban elderly population in China. Hum Vaccin Immunother. 2014;10(10):2994–9. doi:10.4161/21645515.2014.972155

42. National Health Committee of the People’s Republic of China. Yearbook of China Hygiene and Health statistics in 2019. Beijing Union Medical University Press.

