## Supplementary material for "Acceptance of and preference for COVID-19 vaccination in healthcare workers: a comparative analysis and discrete choice experiment": eSupplements

### eSupplement1. Questionnaire used in the survey

Intro:

Thank you for participating in this survey. The research results will provide suggestions for the government to formulate COVID-19 vaccine management policy. The survey involves anonymous filling and does not involve personal privacy, please feel free to answer.

Background:

1. Infection of humans with the new coronavirus (COVID-19) can lead to different health outcomes, including the coronavirus pneumonia.
2. The research and development of COVID-19 vaccines are being actively carried out globally, and some vaccines are about to enter the clinical trial stage to prevent infections caused by COVID-19.
3. In order to enhance the immune protection effect and stability, all vaccines are often added with adjuvants, preservatives and antibiotics allowed by the state during the production process.

Part 1: Demographics information

| Gender | ○ male ○ female |
| --- | --- |
| Education | ○ Master degree and above  ○ Bachelor  ○ College and below |
| Profession | ○ Medical related  ○ others |
| Age group | ○ <20  ○ 20-29  ○ 30-39  ○ 40-49  ○ 50-59  ○ ≥60 |

Part 2: knowledge and attitude of SARS-CoV-2 infection

| Which population is most likely to be infected | ○ the elderly  ○ patients with chronic diseases  ○ infant and child  ○ teenager  ○ adult  ○ all above |
| --- | --- |
| After infection with COVID-19 virus, one might (Multiple choice) | ○ most are asymptomatic or very mild  ○ a few develop into pneumonia  ○ some may be damage to other organs (heart, kidney, etc.)  ○ very few people will die |
| Which population is susceptible to death after being infected with COVID-19 | ○ the elderly and patients with chronic diseases  ○ infant and child  ○ adult  ○ HCWs |
| Are there any effective drugs specifically for the treatment of COVID-19 infection | ○ yes  ○ no  ○ don’t know |
| Will the COVID-19 virus mutate | ○ no  ○ slightly mutation  ○ significant mutation |
| Trend of epidemic in the future | ○ disappear in summer, like SARS  ○ seasonal epidemic, like influenza  ○ nonchronic, like tuberculosis  ○ don’t know |
| Do you think you might be infected with COVID-19 in the future | ○ never  ○ might be  ○ already |

Part 3: the acceptability COVID-19 vaccine

| Do you think it is necessary to develop a COVID-19 vaccine | ○ yes  ○ no  ○ not sure |
| --- | --- |
| How long do you think it will take before the COVID-19 vaccine can be marketed | ○ in 0.5y  ○ in 1y  ○ in 1.5y  ○ longer |
| Who do you think is the priority population for the COVID-19 vaccine (Multiple choice) | ○ HCWs  ○ the elderly  ○ patients with chronic diseases  ○ infant and child  ○ women planning to become pregnant |
| After the National Drug Administration approves the COVID-19 vaccine,  will you be vaccinated | ○ yes  ○ no  ○ not sure |
| If the COVID-19 vaccination needs to be done in stages, how many times can you accept | ○ 1  ○ 2  ○ 3  ○ 4  ○ whatever |
| For you, the minimum protective effect of the vaccine against COVID-19 infection is | ○ <60%  ○ 60-69% (like influenza vaccine)  ○ 70-89%  ○ ≥90% |
| For you, the minimum protective effect of the vaccine against COVID-19 pneumonia is | ○ <60%  ○ 60-69% (like influenza vaccine)  ○ 70-89%  ○ ≥90% |
| Abnormal reactions with a very low probability will occur after vaccination. For you, relative to the severity of pneumonia, which of the following reactions are acceptable  (Multiple choice) | ○ local symptoms such as redness and pain at the injection site  ○ systemic symptoms such as low fever and headache  ○ allergic rashes, etc.  ○ mild liver and kidney damage  ○ serious reactions such as organ damage |
| Are you confident in the future domestic COVID-19 vaccine | ○ very confident, better than foreign vaccines (Skip the next question)  ○ very confident, it’s comparable to foreign vaccines (Skip the next question)  ○ lack of confidence, not good as foreign vaccines  ○ no idea (Skip the next question) |
| The main reason is the domestic vaccine | ○ have low protection effect  ○ poor safety and many adverse reactions  ○ vaccine production is not strict with poor quality  ○ the cold chain of storage and transportation is not strict, made the vaccine is invalid |
| Do you think it is necessary for the country to purchase COVID-19 vaccine to vaccinate the public for free | ○ yes  ○ no |
| If it is at your own expense, what is the highest vaccine pricing you can accept | ○ <100  ○ 100-299  ○ 300-499  ○ >500 |

Part 4: behaviors post epidemic

| After the epidemic, will you take the initiative to vaccinate the following vaccines? | ○ influenza vaccine  ○ PCV13  ○ both  ○ both not |
| --- | --- |
| Will you continue to take these actions after the epidemic is over? | ○ reduce the time to go to crowded places  ○ wash hands frequently  ○ often wear a mask to go out  ○ exercise regularly |

Part5: preferences of vaccination decision

Explanation: Based on the determinants of vaccination, we simulated 9 scenarios, and each scenario was divided into A and B occasions. Please choose the occasion (A or B) according to your judgment.

Situation 1:

| Attributes | occasion A | occasion B |
| --- | --- | --- |
| Infection probability | >30% | 1%-14% |
| Case-fatality ratio | 3%-15% | 3%-15% |
| Trend of epidemic | Disappear in summer, like SARS | Seasonal epidemic, like influenza |
| Vaccine safety | Uncertain, it is a new vaccine | Safe with mild side effects |
| Vaccine effectiveness | 50%-80% | >80% |
| Out of pocket of the vaccination | RMB 300 (US $42.9） | RMB 300 (US $42.9） |
| Acceptance of social contacts | Encourage | Encourage |

Situation 2:

| Attributes | occasion A | occasion B |
| --- | --- | --- |
| Infection probability | 1%-14% | 1%-14% |
| Case-fatality ratio | <3% | 3%-15% |
| Trend of epidemic | Nonchronic, like tuberculosis | Disappear in summer, like SARS |
| Vaccine safety | Uncertain, it is a new vaccine | Safe with mild side effects |
| Vaccine effectiveness | 50%-80% | >80% |
| Out of pocket of the vaccination | RMB 300 (US $42.9） | Free |
| Acceptance of social contacts | Refuse | Encourage |

Situation 3:

| Attributes | occasion A | occasion B |
| --- | --- | --- |
| Infection probability | 1%-14% | 1%-14% |
| Case-fatality ratio | 3%-15% | <3% |
| Trend of epidemic | Seasonal epidemic, like influenza | Nonchronic, like tuberculosis |
| Vaccine safety | Uncertain, it is a new vaccine | Safe with mild side effects |
| Vaccine effectiveness | 50%-80% | 50%-80% |
| Out of pocket of the vaccination | RMB 100 (US $14.3） | RMB 100 (US $14.3） |
| Acceptance of social contacts | Neutral/no opinion | Encourage |

Situation 4:

| Attributes | occasion A | occasion B |
| --- | --- | --- |
| Infection probability | 15%-30% | >30% |
| Case-fatality ratio | <3% | <3% |
| Trend of epidemic | Disappear in summer, like SARS | Seasonal epidemic, like influenza |
| Vaccine safety | Safe with mild side effects | Safe with mild side effects |
| Vaccine effectiveness | 50%-80% | 50%-80% |
| Out of pocket of the vaccination | RMB 300 (US $42.9） | Free |
| Acceptance of social contacts | Neutral/no opinion | Refuse |

Situation 5:

| Attributes | occasion A | occasion B |
| --- | --- | --- |
| Infection probability | 15%-30% | >30% |
| Case-fatality ratio | 3%-15% | 3%-15% |
| Trend of epidemic | Disappear in summer, like SARS | Nonchronic, like tuberculosis |
| Vaccine safety | Safe with mild side effects | Safe with mild side effects |
| Vaccine effectiveness | >80% | >80% |
| Out of pocket of the vaccination | RMB 100 (US $14.3） | Free |
| Acceptance of social contacts | Refuse | Neutral/no opinion |

Situation 6:

| Attributes | occasion A | occasion B |
| --- | --- | --- |
| Infection probability | 1%-14% | 15%-30% |
| Case-fatality ratio | <3% | <3% |
| Trend of epidemic | Disappear in summer, like SARS | Seasonal epidemic, like influenza |
| Vaccine safety | Uncertain, it is a new vaccine | Uncertain, it is a new vaccine |
| Vaccine effectiveness | >80% | >80% |
| Out of pocket of the vaccination | Free | Free |
| Acceptance of social contacts | Neutral/no opinion | Encourage |

Situation 7:

| Attributes | occasion A | occasion B |
| --- | --- | --- |
| Infection probability | >30% | 1%-14% |
| Case-fatality ratio | <3% | 3%-15% |
| Trend of epidemic | Disappear in summer, like SARS | Disappear in summer, like SARS |
| Vaccine safety | Uncertain, it is a new vaccine | Uncertain, it is a new vaccine |
| Vaccine effectiveness | >80% | 50%-80% |
| Out of pocket of the vaccination | RMB 100 (US $14.3） | Free |
| Acceptance of social contacts | Encourage | Refuse |

Situation 8:

| Attributes | occasion A | occasion B |
| --- | --- | --- |
| Infection probability | 1%-14% | 15%-30% |
| Case-fatality ratio | <3% | 3%-15% |
| Trend of epidemic | Disappear in summer, like SARS | Nonchronic, like tuberculosis |
| Vaccine safety | Safe with mild side effects | Uncertain, it is a new vaccine |
| Vaccine effectiveness | 50%-80% | 50%-80% |
| Out of pocket of the vaccination | Free | Free |
| Acceptance of social contacts | Encourage | Encourage |

Situation 9:

| Attributes | occasion A | occasion B |
| --- | --- | --- |
| Infection probability | >30% | 1%-14% |
| Case-fatality ratio | 3%-15% | <3% |
| Trend of epidemic | Seasonal epidemic, like influenza | Disappear in summer, like SARS |
| Vaccine safety | Safe with mild side effects | Uncertain, it is a new vaccine |
| Vaccine effectiveness | >80% | 50%-80% |
| Out of pocket of the vaccination | RMB 100 (US $14.3） | RMB 300 (US $42.9） |
| Acceptance of social contacts | Encourage | Refuse |

**eSupplement2** The attributes in the DCE model

| Attributes | Level of attributes | Evidence |
| --- | --- | --- |
| Probability of infection | 1-14% (reference) | Study of influenza vaccine (PMID: 31186189) , epidemic status worldwide |
|  | 15-30% |  |
|  | >30% |  |
| Severity and probability  of death once infected | <3% (reference) |  |
|  | 3-15% |  |
| Trends of epidemic | Disappear in summer (reference) | Two rounds of Delphi method, based on experts’ opinions |
|  | Seasonal epidemic |  |
|  | Persistent |  |
| Vaccine efficacy | 50-80% (reference) | The 3C model by WHO indicated that the vaccine efficacy and safety were leading contributors in hesitancy; PMID: 28412075 and 20708696 |
|  | >80% |  |
| vaccine safety | Uncertain (reference) |  |
|  | Safe |  |
| Out-of-pocket vaccination cost | Free (reference) | WTP is often measured in DCE model (PMID: 32707831); the influenza vaccine or PPV23 in China |
|  | RMB 100 |  |
|  | RMB 300 |  |
| Social acceptance | Encourage (reference) | Before the vaccine was formerly introduced, different points were noted, which here we used to measure the effect from social contacts |
